# High-Sensitivity Radiation-Free Triage for Adolescent Idiopathic Scoliosis via 3D Point Cloud Geometry

**DOI:** 10.64898/2026.02.11.26346069

**Authors:** Jingfan Yang, Haonan Shi, Zifang Huang, Xiaodong Wang, Wenhui Wang, Tianyuan Zhang, Jingwen Wang, Yifan Zhan, Hui Liu, Zifang Zhang, Jiannan Zhang, Zhijun Fei, Xiaoling Xuan, Yihao Gao, Yaolong Deng, Liming Wang, Xiyang Liu, Long Tian, Yaqing Zhang, Lirong Ai, Junlin Yangx

## Abstract

Widespread screening for Adolescent Idiopathic Scoliosis (AIS) is critical for early intervention, yet it is currently bottlenecked by the inherent limitations of traditional methods. Radiographic diagnosis poses cumulative radiation risks, while manual physical examinations are highly subjective and time-consuming. Recent non-invasive 2D computer vision approaches suffer from an unavoidable “dimensionality gap,” failing to capture critical depth and rotational information, which frequently leads to diagnostic misjudgments. To address these challenges, we present PointScol, a high-sensitivity, radiation-free triage system leveraging direct geometric processing of 3D back surface point clouds. Our framework employs a sequential pipeline: first, an automated segmentation module rigorously standardizes the input geometry by isolating the dorsal region of interest; subsequently, a diagnostic classification module evaluates the spinal deformity. Validation on a multi-center dataset (n=128) demonstrated that for the primary screening task (10° Cobb angle threshold), PointScol achieved 100.00% sensitivity in the external cohort, acting as a reliable gatekeeper to safely rule out healthy individuals without missing any cases requiring referral. Building upon the robust accuracy established at this 10° baseline, an extended 5-class grading module provides further diagnostic value. Rather than functioning as a rigid predictive task, this multi-class stratification acts as an advanced clinical assistant, offering nuanced severity insights to guide referral urgency and optimize medical resource allocation for high-risk patients. Collectively, this sequential design establishes PointScol as a safe and highly efficient clinical filter: it reliably prevents unnecessary radiation exposure for healthy adolescents while ensuring prioritized interventions for those most in need.

## Introduction

Adolescent Idiopathic Scoliosis (AIS) is a complex three-dimensional spinal deformity characterized by a lateral curvature with a Cobb angle greater than 10°. Affecting 3% to 5% of adolescents, the condition can lead to severe thoracic asymmetry, cardiopulmonary dysfunction, and psychological distress if left untreated.^1,2^ Consequently, widespread screening is critical to ensure timely intervention, preventing disease progression and improving long-term quality of life.^3^ Optimal screening requires a delicate balance.^4^ High sensitivity is critical to identify all potential cases for referral—particularly at the 10° Cobb angle threshold, where early detection allows for proactive, radiation-free longitudinal observation and follow-up rather than immediate intervention. Simultaneously, in the context of large-scale adolescent population screening, high specificity is paramount to prevent the over-referral of healthy individuals, thereby minimizing unnecessary cumulative radiation exposure, mitigating psychological anxiety, and avoiding the depletion of specialized medical resources. While recent 2D visible-light (RGB) screening approaches offer non-invasive alternatives, they inherently suffer from a “dimensionality gap,” lacking the depth and rotational information necessary to accurately rule out false positives. By directly capturing true 3D surface geometry, our approach overcomes these planar limitations, demonstrating that superior topological imaging fundamentally drives the high specificity and diagnostic fidelity required for reliable mass-scale triage.

While the radiographic Cobb angle remains the clinical “gold standard” for diagnosing scoliosis, its reliance on ionizing radiation poses significant cumulative health risks, particularly for adolescents requiring serial monitoring.^5^ Although radiation-free alternatives such as freehand 3D ultrasound have emerged, their widespread deployment is often constrained by high operator dependency, the need for complex tracking equipment, and low clinical throughput.^6,7^ To address these bottlenecks, recent computer vision approaches have leveraged deep learning to analyze 2D back surface photography or synthesize virtual X-rays via generative adversarial networks (GANs).^8,9^ However, scoliosis is inherently a complex, three-dimensional volumetric deformity. Purely 2D-based models suffer from an unavoidable “dimensionality gap,” failing to capture critical rotational depth information. Furthermore, generative approaches in the absence of geometric constraints are prone to “hallucinations,” introducing non-existent anatomical features that compromise diagnostic reliability.^9^

To overcome these planar limitations, research paradigms have pivoted towards capturing volumetric data via markerless surface topography and RGB-D sensing,^10,11^ alongside dynamic analysis of gait sequences.^12^ The theoretical superiority of direct 3D learning is well-established; point cloud architectures are capable of extracting local geometric features of asymmetry that are unattainable by 2D methods.^13^ Despite this technological maturity, a critical gap remains in clinical translation: existing 3D tools primarily focus on shape reconstruction or scalar regression of the Cobb angle.^14^ There is a notable lack of dedicated frameworks designed specifically as high-sensitivity triage filters—tools capable of safely “ruling out” healthy individuals.^15^ Moreover, current 3D deep learning models often operate as “black boxes,” lacking the geometric interpretability necessary to foster clinical trust.^16^

Herein, we present PointScol, a novel radiation-free triage system that reconciles screening safety with diagnostic granularity, with its overall workflow and system architecture illustrated in Fig. 1. Our framework employs a streamlined two-stage, dual-task deep learning architecture. First, to ensure data fidelity, an automated segmentation module isolates the dorsal region of interest (ROI), eliminating environmental noise that plagues traditional non-contact screenings. Second, the system executes a hierarchical diagnostic protocol: (1) A Binary Screening Module functions as a high-sensitivity gatekeeper to identify all potential risk cases (>10°); (2) A Fine-grained Grading Module then stratifies these cases into five severity classes. This design allows the system to achieve “zero-miss” safety for exclusion while providing specific severity contexts (e.g., distinguishing observation candidates from surgical urgencies) to guide referral urgency. Developed and validated on a large-scale, multi-center dataset comprising 776 individuals with paired 3D scans and X-ray ground truths, our system demonstrates distinct utility across these tasks. PointScol achieves a sensitivity of 100.00% for the primary screening threshold, effectively ensuring no false negatives. Simultaneously, its severity grading capability offers a reliable “second opinion” for resource allocation, bridging the gap between automated detection and personalized patient management.

**Figure 1.**
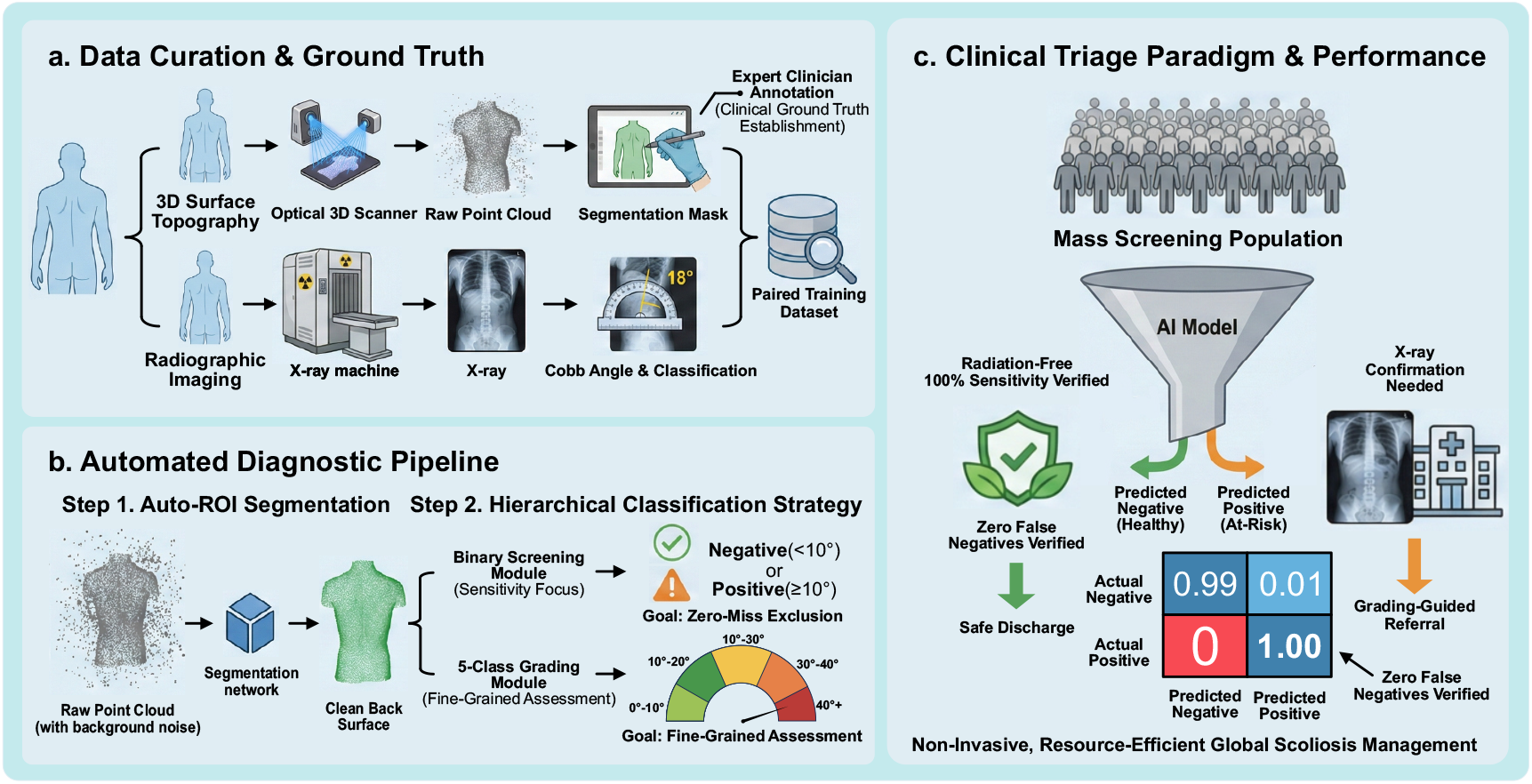
Overview of the radiation-free AIS screening framework. (a) Data Curation and Ground Truth Establishment. A multi-modal dataset is constructed by pairing radiation-free 3D surface topography with radiographic (X-ray) ground truth, annotated by expert clinicians. Automated Diagnostic Pipeline. The framework consists of an automated ROI segmentation module for data standardization, followed by a hierarchical classification strategy. A Binary Screening Module first prioritizes high sensitivity to ensure safety, while a subsequent 5-Class Grading Module provides fine-grained severity assessment for individuals identified as at-risk. Clinical Triage Paradigm and Performance. The system operates as a “zero-miss” filter to safely discharge healthy individuals (0% false-negative rate) and stratifies at-risk patients to guide referral urgency, thereby maximizing clinical resource efficiency in mass screening.

The primary contributions of this work include:

- **Establishment of a Multi-Center 3D Benchmark:** We constructed a comprehensive, large-scale dataset comprising 776 individuals across three independent institutions. By rigorously pairing 3D back surface point clouds with X-ray ground truths, we addressed the critical scarcity of high-quality, annotated 3D data in orthopedic AI.
- **A Sequential “Safety-First” Triage Paradigm:** We propose a novel workflow that combines high-fidelity ROI extraction with a dual-objective diagnosis. This sequential structure guarantees near-zero false negatives (Sensitivity: 100%) for safe exclusion while offering clinically actionable severity stratification (5-class grading) to optimize referral efficiency.
- **Interpretability and Clinical Trustworthiness:** By incorporating geometric feature visualization, our system bridges the gap between “black box” AI predictions and clinical decision-making.

## Results

### Overview of datasets

To develop and validate the radiation-free scoliosis screening system, we established an internal Development Cohort and an independent Multi-Center Validation Cohort. For the Development Cohort, we retrospectively analyzed data from 648 participants who underwent high-resolution 3D back surface topological scanning and matched full-spine radiography at Xinhua Hospital (Shanghai Jiao Tong University School of Medicine) between 2022 and 2024. Patients were included strictly if they had no prior spinal surgery and underwent same-day acquisition of 3D back surface data and radiographic imaging. Each patient contributed a single data sample to ensure independence and prevent data leakage. We randomly allocated 80% of these cases to the training set (n = 519) for model optimization and the remaining 20% to the internal testing set (n = 129).

Moreover, to explicitly assess the system’s robustness against scanner variability, population shifts, and operational biases, we established the Multi-Center Validation Cohort comprising 128 individuals. These patients were recruited from two distinct external institutions (The Third Affiliated Hospital of Sun Yat-sen University and Xin Miao Scoliosis Treatment Centre) between October 2022 and September 2024. Data acquisition for this cohort followed the exact same inclusion criteria but utilized different operator teams and environmental setups.

To ensure that structural variations in the 3D surface data reflect objective human diversity rather than operational artifacts or single-center bias, we detailed the anthropometric distributions across both datasets. The Development Cohort reflected a broad natural distribution: a mean age of 13.56 ± 2.96 years (range: 8–18), mean height of 159.67 ± 11.04 cm, mean weight of 47.13 ± 10.47 kg, and a Body Mass Index (BMI) ranging from 11.05 to 34.69 kg/m^2^ (mean: 18.33 ± 2.98). Similarly, the Multi-Center Validation Cohort demonstrated consistent natural epidemiological variations, with a mean age of 13.24 ± 2.11 years (range: 7–18), mean height of 158.24 ± 10.13 cm, mean weight of 46.34 ± 10.50 kg, and BMI ranging from 12.40 to 34.16 kg/m^2^ (mean: 18.37 ± 3.31). This extensive and naturally occurring morphological variation across independent clinical centers guarantees that our network learns robust, intrinsic anatomical features resilient to soft tissue differences, rather than overfitting to cohort-specific noise.

### Automated ROI Segmentation and Preprocessing

A critical prerequisite for high-throughput, reproducible screening is the fully automated isolation of the dorsal Region-of-Interest (ROI) from raw 3D scans. Raw acquisition data invariably contain significant environmental noise (e.g., background artifacts) and non-relevant biological structures that can confound downstream diagnostic algorithms. To address this, we implemented a segmentation framework, an attention-based network selected for its ability to capture local geometric details within global shape contexts.

Performance evaluation was stratified across an internal test set and an independent external test set to verify algorithmic robustness (Supplementary Table 2). On the internal test set, the model demonstrated high segmentation fidelity, achieving a Point Accuracy (PA) of 92.57% (95% CI: 91.48%–93.66%) and a dorsal Intersection over Union (IOU) of 89.70% (95% CI: 88.17%–91.23%). Crucially, to assess clinical generalizability, we validated the pipeline on the external test set. The model maintained consistent performance with a PA of 91.83% (95% CI: 91.02%–92.64%) and a dorsal IOU of 87.89% (95% CI: 87.20%–88.58%), indicating minimal performance degradation across distinct data sources.

To validate the clinical applicability of these quantitative results, we conducted a qualitative assessment of segmentation fidelity (Supplementary Fig. 1). Visualization of the IOU distribution and error maps reveals that the pipeline precisely delineates anatomical boundaries—including the shoulders, waistline, and iliac crests. The method demonstrated high stability across subjects with varying morphological characteristics, effectively handling sparse point density and surface curvature, largely reducing the need for manual intervention. This high-fidelity segmentation ensures that the subsequent diagnostic module receives standardized, highly clean topological input, decoupling diagnostic accuracy from variations in patient positioning or background clutter.

### Binary Classification Performance and Clinical Generalizability

To operationalize the screening task, we defined the binary classification criteria as scoliosis (Cobb angle >10°) versus non-scoliosis (0°–10°). Upon internal validation, the deep learning framework demonstrated high discriminative fidelity. As illustrated in the normalized confusion matrices (Fig. 2a), the soft voting ensemble strategy correctly identified all positive and negative samples, resulting in an accuracy of 100% and a corresponding Area Under the Curve (AUC) of 1.00 (Fig. 3a). To further elucidate the ensemble’s efficacy, microscopic error analysis on challenging instances (Fig.2b) revealed strong error complementarity among the individual expert models. The soft voting mechanism successfully leveraged these divergent probabilities to correct individual misclassifications. Furthermore, analysis of straightforward cases (Fig. 2c) demonstrated that the individual models maintained consistently high correct-label prediction probabilities, providing a highly confident baseline. This flawless internal performance underscores the inherent diagnostic advantages of 3D point cloud imaging; by directly capturing spatial depth and rotational asymmetry, the modality provides comprehensive geometric features that successfully bypass the dimensionality loss typical of 2D approaches. Consequently, the model effectively extracted the distinct morphological signatures of spinal deformities without overfitting, establishing a robust baseline for subsequent external testing.

**Figure 2.**
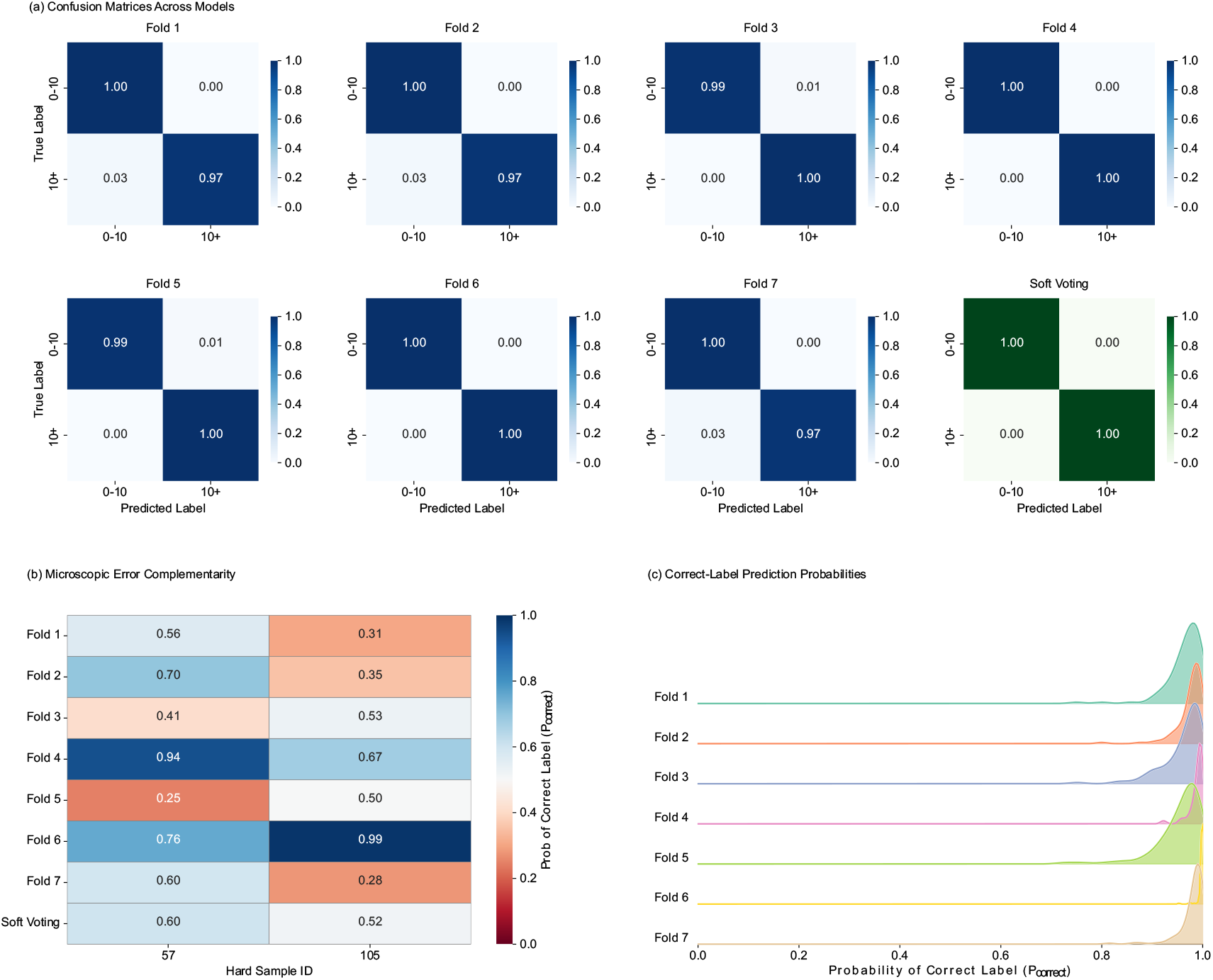
Classification performance and prediction probability distributions on the internal test dataset. a, Normalized confusion matrices for the binary classification (‘0°–10°’ and ‘10°+’) across the seven individual models (Folds 1–7; blue scale) and the final soft voting ensemble (green scale). Diagonal elements represent the proportion of correct predictions. b, Heatmap illustrating microscopic error complementarity for identified hard samples. Cell values represent the probability of predicting the correct label (*P*_*correct*_) by each fold and the soft voting mechanism. A diverging color scale distinguishes correct (*P*_*correct*_ > 0.5, blue) from incorrect (*P*_*correct*_ < 0.5, red) predictions. c, Ridge plot showing the kernel density estimation of *P*_*correct*_ across the seven folds for straightforward cases (easy samples with no model errors).

**Figure 3.**
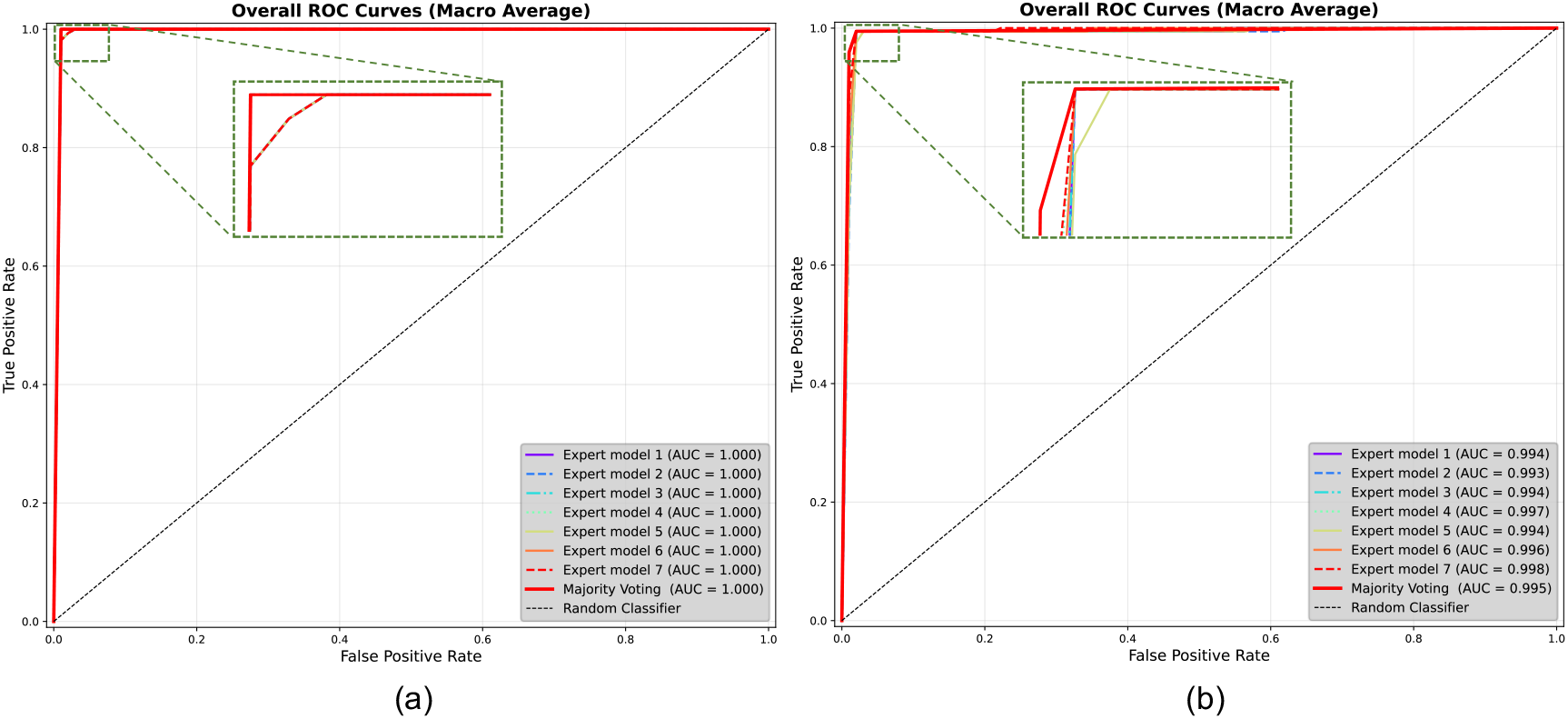
Performance evaluation of expert models and ensemble voting strategy in binary classification. a, b, Macro-average Receiver Operating Characteristic (ROC) curves derived from the internal testing set (a) and the external testing set (b). The plots compare the classification performance of seven individual expert models (colored dashed lines) against the voting ensemble model (solid red line). The insets (green dashed boxes) provide magnified views of the top-left corner to highlight performance distinctions in the high-sensitivity and high-specificity regions. Area Under the Curve (AUC) values for each model are listed in the legend, demonstrating the robustness of the ensemble approach across both datasets.

To rigorously evaluate the model’s utility in a real-world setting, we deployed the trained ensemble on an independent external validation set. The model exhibited strong generalization capabilities, maintaining a high classification accuracy of 99.24% (95% CI: 97.66%–100.00%). The Receiver Operating Characteristic (ROC) analysis further confirmed this robustness, yielding an AUC of 99.54% (95% CI: 98.32%–100.00%) (Fig. 3b). Notably, the soft voting ensemble—which aggregates prediction probabilities from seven independent expert models—proved superior to individual models. By effectively exploiting microscopic error complementarity (Fig. 4b), it mitigated variance and stabilizing the decision boundary against data heterogeneity inherent in multi-center datasets (e.g., variations in hardware calibration, scanning environments, and patient demographics) (Fig. 3b).

**Figure 4.**
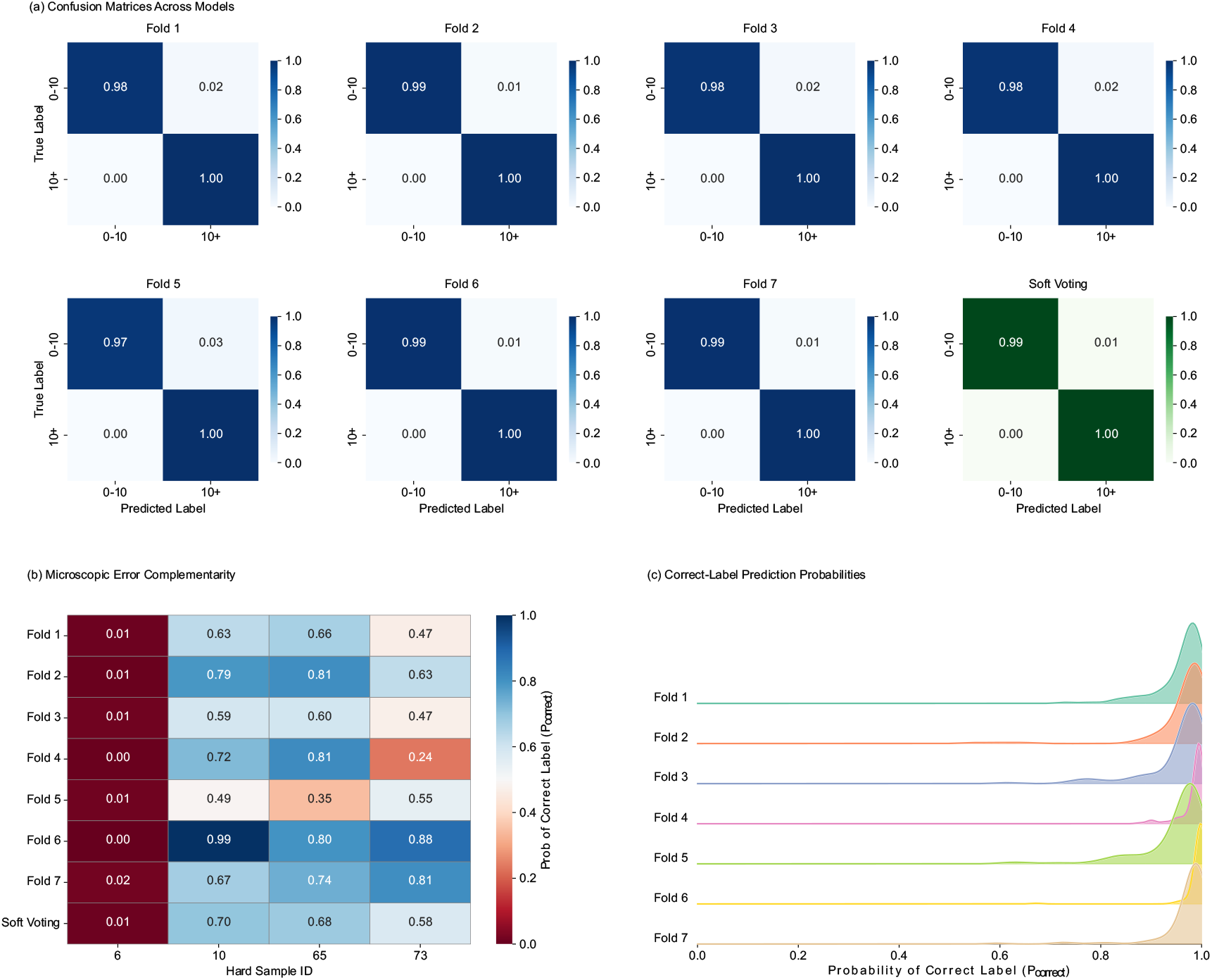
Generalization assessment and prediction probability distributions on the multi-center validation cohort. a, Normalized confusion matrices evaluating the predictive performance of the seven individual models (Folds 1–7; blue scale) and the soft voting ensemble (green scale) on unseen external data. Diagonal elements denote the proportion of correct classifications. b, Heatmap of microscopic error complementarity for hard samples within the external cohort, displaying *P*_*correct*_ for each model and the ensemble using a diverging color scale. c, Ridge plot detailing the kernel density estimation of *P*_*correct*_ across the individual models for easy samples in the external dataset.

Crucially for a triage system, PointScol achieved 100.00% sensitivity (recall) and a 100.00% Negative Predictive Value (NPV) in the external cohort (Supplementary Table 3), successfully identifying all patients with a Cobb angle >10°. Accompanying this perfect sensitivity, the model maintained a high specificity of 98.93% (95% CI: 96.33%–100.00%) and a Positive Predictive Value (PPV) of 97.40%. Error analysis via confusion matrices (Fig. 4a) confirmed only a single false-positive instance across the entire external dataset, while the probability distributions demonstrated sustained diagnostic confidence even on unseen straightforward cases (Fig. 4c).

Comprehensive statistical evaluation further underscores the reliability of this 3D-geometric approach. The F1-score, harmonizing precision and recall, reached 98.67% for the external set, while the Matthews Correlation Coefficient (MCC) was 98.16%, indicating balanced predictive performance robust to potential class imbalances (Supplementary Table 3). The consistency observed between the individual expert models and the final voting outcome (Figs. 2, 3 and 4) confirms that the proposed point cloud analysis pipeline provides a stable, reproducible, and highly accurate solution for automated scoliosis screening. Ultimately, by achieving a zero-miss rate (100.00% NPV), PointScol fulfills the core mandate of mass screening: it acts as a highly reliable, radiation-free clinical gatekeeper. This ensures that healthy adolescents are safely ruled out from unnecessary radiographic exposure, while all at-risk individuals are accurately flagged for prioritized clinical intervention.

### Fine-grained Severity Stratification and Clinical Grading

To advance the framework’s utility from initial screening to precise clinical grading, we extended the evaluation to a fine-grained five-grade stratification task (Supplementary Table 4). Crucially, even under increased classification complexity, the model preserved the robust identification of non-scoliosis cases (0°–10°) established in the binary baseline (Supplementary Fig. 2). While the exact binning of intermediate cases remains inherently challenging due to the continuous morphological progression of spinal deformation, the model’s errors were predominantly confined to immediately adjacent severity grades rather than catastrophic outlier misclassifications. Additionally, although cases of extreme severity (>40°) were naturally scarce in the cohort, the model maintained a robust capacity to identify severe progressions and rule out false positives across both internal and external datasets (Supplementary Figs. 3 and 4).

To further validate the model’s utility in clinical decision-making, we derived binary performance metrics based on the 5-class probability distributions at critical intervention thresholds: Cobb angles of 10° (screening), 20° (observation vs. bracing), and 40° (bracing vs. surgery) (Supplementary Fig. 5, Supplementary Table 5). For the 20° threshold, which encompasses a more substantial portion of the clinical cohort, we evaluated the practical trade-off in bracing decisions using operational metrics. In contrast, for the critical 40° surgical threshold—where positive cases are inherently rare and sample sizes are limited—we utilized the area under the curve (AUC) to assess the model’s overall discriminatory capability, rather than relying on threshold-dependent metrics that are highly susceptible to small sample fluctuations. These results indicate that the fine-grained feature representation can be flexibly adapted to various clinical cutoff requirements. Across both internal testing and the independent external dataset (Supplementary Fig. 6), the system effectively acts as a reliable “second opinion”—anchoring the healthy baseline while successfully flagging potential severe cases for prioritized specialist review.

### Interpretability and Anatomical Alignment

To foster clinical trust and validate that the classification module leverages biologically relevant features rather than spurious correlations (e.g., background artifacts or clothing), we visualized the model’s decision-making process using point-wise saliency mapping. As illustrated in Fig. 5, we present a comparative visualization of the input 3D point cloud back surface alongside the corresponding ground-truth X-ray radiograph.

**Figure 5.**
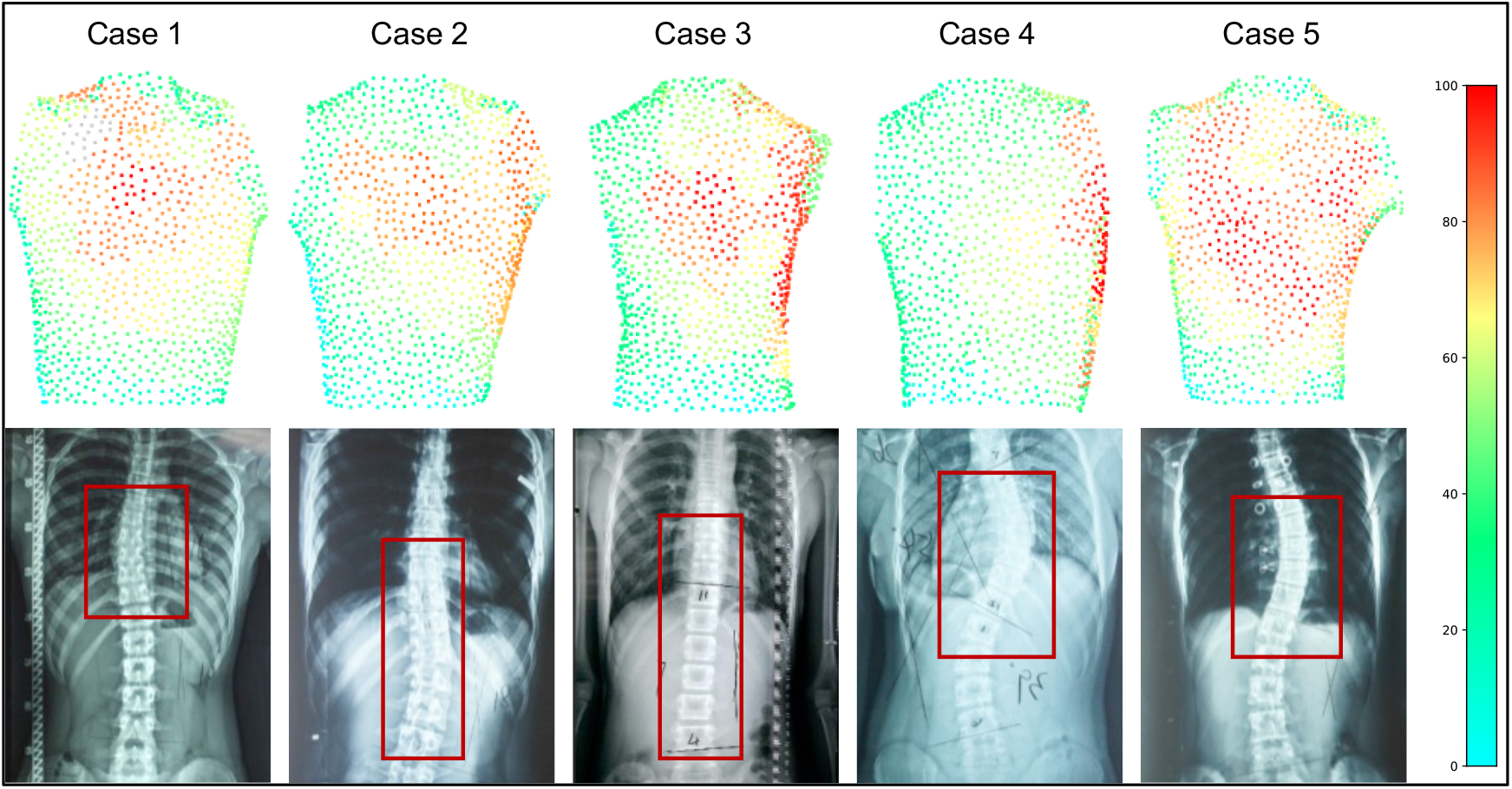
Visualization of model attention regions compared to anatomical ground truth. Heatmap visualizations of attention regions for the classification model are presented. The first row displays the point cloud attention maps generated by the model, where warmer colors (red) indicate higher model attention. The second row shows the corresponding X-ray images with clinically annotated scoliosis regions.

The attention heatmaps reveal that the model consistently assigns high importance weights to paravertebral regions exhibiting pathological asymmetry, effectively filtering out irrelevant peripheral areas. Specifically, the model’s focus is concentrated on key orthopedic markers: the scapular prominence, the lateral waistline asymmetry, and the paraspinal rib hump. Overlaying these attention maps with the spinal radiographs demonstrates a high degree of spatial alignment between the AI-highlighted surface regions and the underlying skeletal deformity. The “hotspots” on the 3D surface correspond closely with the spinal curvature and the convex side of the rib cage visible on the X-ray.

This qualitative alignment is clinically significant, as the rib hump is the direct topological manifestation of vertebral rotation—a critical biomechanical driver of scoliosis progression. By focusing on these anatomical indicators rather than global image statistics, the classification module provides clinicians with a visual rationale alongside the risk prediction, thereby supporting informed referral decisions based on established markers of deformity.

## Discussion

The clinical management of adolescent idiopathic scoliosis (AIS) has historically been trapped in a reactionary cycle, paralyzed by a critical conflict: the imperative for early detection versus the carcinogenic risks of cumulative radiation exposure. Current screening pathways are bottlenecked by reliance on subjective physical examinations (e.g., the Adam’s Forward Bend Test), which suffer from high inter-rater variability and frequently result in missed early-stage diagnoses.^17^ Conversely, definitive radiographic confirmation imposes unnecessary radiation burdens on healthy adolescents.^18^ To resolve this historic impasse, we present a radiation-free, fully automated screening framework that transforms 3D back surface point clouds into objective diagnostic biomarkers. By bridging the gap between safety and accuracy, our work shifts the clinical paradigm from intermittent, reactive diagnosis based on visible deformity to continuous, proactive surveillance.

Our study makes three distinct contributions to the field of orthopedic AI. Our findings primarily establish the system’s utility as a high-reliability clinical gatekeeper. Aligning with the urgent need for high-sensitivity triage, PointScol achieved 100% sensitivity at the primary screening threshold of 10° across multi-center cohorts. Clinically, while a 10° curvature typically requires active observation rather than immediate intervention, it strictly mandates rigorous longitudinal follow-up. Because our imaging method is entirely radiation-free, it uniquely enables large-scale, early-stage, and continuous monitoring for these borderline cases without the compounding carcinogenic risks of X-rays. This operationalizes a “near-zero false-negative” strategy, ensuring that healthy individuals are safely excluded from the referral pipeline without radiographic confirmation. In healthcare systems where cost-efficiency is paramount, this automated, low-cost triage significantly reduces the economic burden of unnecessary X-rays and specialist consultations, while simultaneously alleviating the substantial psychological distress individuals and families experience during “false alarm” referrals.

Methodologically, the high diagnostic performance of PointScol is intrinsically driven by the inherent advantages of the 3D surface imaging modality itself. Our approach addresses the limitations of previous computer vision techniques by prioritizing geometric fidelity over texture. Unlike 2D photographic models that suffer from the ‘dimensionality gap’ and are easily confounded by lighting or skin texture, our direct 3D processing captures the intrinsic topological asymmetry essential for volumetric analysis. Furthermore, as a highly cost-effective and low-resource acquisition method, 3D surface scanning serves as an ideal structural supplement to conventional 2D screening, offering vastly superior spatial information at a fraction of the cost of radiography. This strategy dovetails with a growing consensus in high-precision computational medicine: that direct geometric processing—operating on raw point clouds or surface meshes—far surpasses voxel-based or multi-view approaches in resolving complex topologies. A prime example lies in digital dentistry, where geometric deep learning networks (e.g., MeshSegNet) have successfully replaced manual segmentation by directly analyzing the curvature and normal vectors of intraoral scan data.^19^ Similarly, in vascular diagnostics, point-based architectures like PointNet++ are now deployed to map coronary centerlines with sub-millimeter precision, bypassing the resolution loss inherent in volumetric grids.^20^ By translating these geometric principles to the spinal dorsal surface, our PointScol framework confirms that raw coordinate data contains the richest diagnostic signal, uncorrupted by projection artifacts.

We address the “black box” skepticism that limits the adoption of AI in specialized diagnostics.^21^ Prioritizing clinical interpretability alongside accuracy, our system incorporates geometric feature visualization to reveal the specific regions of asymmetry driving the diagnosis. This transparency fosters the clinical trust necessary for medical integration. Additionally, we validated our model on a large-scale, multi-center benchmark of paired 3D and X-ray data, confirming that clinically actionable insights can be derived from non-invasive morphological data alone, offering a richer information profile than previous alternatives.

Despite these advancements, several limitations warrant careful consideration. While the proposed five-category classification framework is intended to provide granular risk stratification, its effectiveness is inherently dependent on adequate and balanced data coverage across the full Cobb angle spectrum. In the present dataset, uneven representation within certain angle intervals restricts the model’s ability to achieve uniformly high performance across all categories. Nevertheless, from a clinical standpoint, the primary objective remains the reliable binary discrimination between normal and pathological cases, a task for which our model demonstrated consistently strong performance. Building on this foundation, future work will prioritize expanding and balancing the training dataset across underrepresented Cobb angle ranges to further improve fine-grained multi-category classification.

A key limitation is that the dataset was derived exclusively from a Chinese population, which may limit generalizability given potential ethnic differences in BMI distribution and trunk morphology. Moreover, despite the use of a multicenter cohort, the overall sample size remains limited, and further expansion across additional centers and populations will be necessary to more rigorously assess the robustness and generalizability of the proposed approach in large-scale prospective studies.

The radiation-free design allows safe longitudinal disease monitoring, addressing the clinical need for frequent assessments during growth-related progression in adolescent scoliosis. By mitigating the risks associated with cumulative radiographic exposure, this approach not only enhances patient safety but also supports a paradigm shift toward more accessible, non-invasive screening strategies, with profound implications for health equity in scoliosis management.

## Methods

### Data Cohorts and Preprocessing

This study was approved by the Ethics Committee of Xinhua Hospital Affiliated to Shanghai Jiao Tong University School of Medicine and conducted in accordance with the Declaration of Helsinki. Due to the retrospective nature of the development dataset, the requirement for informed consent was waived, whereas written informed consent was obtained from all participants in the external validation cohort. The dataset utilized in this study was collaboratively collected over a comprehensive two-year period, from October 2022 to September 2024, involving a primary development center and two independent validation centers. To simulate a resource-constrained screening environment, all 3D back surface topographical data were acquired using a portable, consumer-grade 3D scanning solution integrated with a mobile interface. This specific hardware configuration was selected to demonstrate the feasibility of low-cost, radiation-free mass screening. To establish the radiological ground truth for spinal deformity, standing full-spine high-resolution X-ray imaging was acquired synchronously with the point cloud scans for all subjects. The Cobb angle, serving as the gold standard for diagnosing and grading scoliosis, was measured and verified by senior radiologists to ensure the reliability of the labels used for model development.

The study design incorporated two geographically and institutionally distinct cohorts to rigorously evaluate model robustness against real-world variations. The Xinhua Development Cohort served as the foundational dataset for model training and internal parameter optimization. This cohort contributed a total of 648 samples collected under controlled clinical conditions. To ensure the model’s generalizability and robustness, we established an independent Multi-Center Validation Cohort, comprising 128 samples collected from The Third Affiliated Hospital of Sun Yat-sen University and the Xin Miao Scoliosis Treatment Centre. This multi-center approach was explicitly designed to expose the model to heterogeneity in patient demographics, slight variations in imaging instrumentation, and procedural nuances inherent to different clinical settings. Strict inclusion and exclusion criteria were applied uniformly across all centers: participants were excluded if they had a history of spinal surgery, visible physical defects unrelated to scoliosis that might distort back topology, or incomplete imaging data. To prevent data leakage and ensure an unbiased evaluation, the dataset was partitioned strictly on a per-patient basis.

To facilitate high-throughput mass screening and eliminate inter-operator variability, we implemented a fully automated preprocessing pipeline to normalize raw scan data for the downstream classification network (Supplementary Fig. 7). First, raw 3D data acquired in stereolithography (STL) format were converted into point cloud representations via point sampling. To isolate the Region of Interest (ROI), we employed a deep learning segmentation network to automatically extract the posterior torso (lower neck, trunk, and lower back) while computationally excising the head and limbs. Ground truth for this network was established by clinical experts, who manually annotated the anatomical ROI using standard 3D point cloud processing software.

Following segmentation, the raw point clouds exhibited varying densities (distributions for Training, Internal Test, and External Test sets are shown in Supplementary Fig. 8). To standardize input resolution, all samples were normalized to a fixed target resolution. High-density scans were downsampled using standardized point sampling techniques to maximally preserve geometric topology, while sparse scans were upsampled via interpolation methods. This normalization ensures that feature density remains invariant across individuals of different body sizes, directing the classifier to focus on topographical shape asymmetries indicative of scoliosis rather than artifacts arising from scanning variations.

### ROI Extraction via 3D Point Cloud Segmentation

To precisely isolate the dorsal Region of Interest (ROI) from raw back surface scans—a critical step to eliminate environmental noise and clothing artifacts—we employed an attention-based point cloud segmentation network. Unlike projection-based or voxel-based methods that rasterize data into regular grids, often causing a loss of fine-grained topological details, our approach operates directly on the unordered set of points in their native continuous metric space. This is particularly advantageous for preserving the intrinsic surface curvature and subtle asymmetry required for scoliosis assessment.

The network employs an encoder-decoder architecture adapted for dense semantic segmentation, utilizing a self-attention mechanism. Specifically, for a local neighborhood of points, the model captures complex local geometric relationships such as the scapular prominence and the lateral waistline rather than relying solely on global shape statistics. The encoder progressively reduces point cardinality while increasing feature dimensionality, ensuring robust feature pooling even across scans with varying point densities. The symmetric decoder then recovers the high-resolution point set, fusing deep semantic features with shallow geometric details to accurately delineate anatomical boundaries along the neck and lower back.

The selection of this architecture offers distinct advantages for processing biological surface topology in a clinical screening context. First, the attention operator is permutation-invariant, making it mathematically robust to the irregular and unstructured nature of handheld 3D scans. Second, the model achieves adaptive modulation of feature channels, which significantly enhances its ability to differentiate smooth skin surfaces from background noise. Third, the integration of geometric contextual encoding enables the network to learn relative spatial relationships, which is critical for recognizing the standardized dorsal orientation required for subsequent deformity analysis. Optimization was performed using standard gradient descent methodologies with customized learning rate scheduling to ensure convergence.

### Classification Model

To efficiently adapt pre-trained point cloud models to the specific morphological nuances of AIS, we employed a parameter-efficient fine-tuning framework. Unlike standard fully fine-tuning (FFT) strategies that update all model parameters—often leading to overfitting on limited clinical datasets—this approach freezes the pre-trained Transformer backbone and integrates a lightweight, trainable spectral adapter module into the network layers. We selected this spectral architecture specifically for its ability to decompose complex spinal deformities into distinct geometric frequencies: identifying global curvature trends while isolating subtle local asymmetries (e.g., rib humps) that are often obscured in the spatial domain.

The core mechanism operates by transforming input tokens from the spatial domain to the spectral domain using the Graph Fourier Transform (GFT). Specifically, tokens are down-projected to generate a low-rank graph signal. We then apply the GFT by multiplying this signal with pre-computed spectral bases, effectively decomposing the spatial signals into orthogonal frequency components. This step is clinically pivotal, as it effectively decouples the holistic torso shape (low-frequency components) from pathological local protrusions (high-frequency components). In the spectral domain, a specialized adaptation layer modifies these spectral coefficients. This process allows for targeted tuning by modifying the coefficients within the eigenvector space. Finally, an inverse GFT restores the features to the spatial domain, which are then integrated back into the computation flow.

To enable robust spectral analysis of the dorsal topography, we constructed multi-scale point cloud graphs, comprising a global graph to capture the overall spinal alignment and localized sub-graphs to resolve fine-grained paraspinal details. Edge weights for these graphs were defined using a distance-based scaling strategy. We computed the corresponding Laplacian matrices and performed eigenvalue decomposition to obtain orthogonal eigenvector matrices, ensuring that the intrinsic geometric information of our specific point clouds was explicitly incorporated into the fine-tuning process. The model was trained using a standard optimizer with weight decay, coupled with a cosine learning rate scheduler.

### Training Strategy and Ensemble Inference

To ensure robust model performance and mitigate overfitting, distinct training strategies were adopted for the segmentation and classification tasks. For the segmentation network, a fixed data partitioning scheme was applied, dividing the dataset into a training set, an internal held-out test set, and an independent external multi-center test set. To establish high-quality ground truth, all segmentation masks underwent a secondary review by senior radiologists. This process involved manual refinement of samples in which automated thresholding failed due to factors such as hair occlusion or clothing artifacts, ensuring anatomically accurate boundary annotations for model learning.

For the classification task, an ensemble learning strategy based on random subsampling was employed to enhance model robustness. The development cohort was used to generate multiple distinct expert models, each trained using a random split of 60% for training and 40% for internal validation. The number of expert models was empirically chosen to ensure comprehensive data coverage and model diversity, effectively stabilizing the predictive variance without incurring excessive computational overhead. Importantly, the multi-center validation cohort was kept completely isolated throughout training and hyperparameter optimization and was accessed only once for final performance evaluation, thereby providing a stringent assessment of previously unseen populations.

During the inference stage, a soft-voting ensemble strategy was applied. Instead of outputting discrete class labels, each expert model generated independent prediction probabilities. The final aggregated probability for a given case was calculated by averaging these probabilities across all models, and the ultimate diagnostic decision was derived from this consensus probability. This continuous aggregation approach preserves the confidence gradients necessary for threshold-independent evaluations (e.g., AUC curves), effectively reduces prediction variance from individual models, and yields a more stable and reliable screening decision.

### Deciphering morphological signatures

To provide clinical interpretability and visualize the morphological features driving the AI’s decision, we generated class activation heatmaps based on our Transformer-based architecture. Unlike standard point-wise networks, our model processes the input point cloud by grouping local points into patches and utilizing a self-attention mechanism. To create the visualization, we extracted the self-attention maps from the final layer of the Transformer encoder. Specifically, we calculated the attention weights between the global class token and each local patch token. These weights quantify the contribution of each local geometric structure to the global shape descriptor. Since the model operates on grouped points, the patch-level attention scores were interpolated and mapped back to the original 3D spatial coordinates of the constituent points.

### Statistical Analysis

Patient-level screening assessments were derived from the aggregated output of the ensemble deep learning framework. To ensure robust classification, we integrated predictions from independent expert models. Specifically, the raw confidence scores (logits) from each model were normalized into probability distributions via a softmax function. The final aggregated probability for each participant was determined using a soft-voting strategy. The ultimate classification decision was then made by selecting the class with the maximum aggregated probability. For the binary screening task, participants were classified as positive for scoliosis when this aggregated probability for the positive class exceeded the optimal decision threshold derived from the internal development set.

In evaluating the system’s overall classification performance, we calculated a comprehensive suite of diagnostic metrics, including Accuracy, Sensitivity, Specificity, Positive Predictive Value (PPV), Negative Predictive Value (NPV), F1-score, and the Matthews Correlation Coefficient (MCC). The threshold-independent discriminative capability of the model was further assessed using the Area Under the Receiver Operating Characteristic curve (AUC) and Average Precision (AP).

For the automated ROI segmentation module, geometric fidelity and point-level congruence were quantified using Point Accuracy (PA), Class Point Accuracy (CPA), Intersection over Union (IoU), and mean Intersection over Union (mIoU). Given the unique spatial characteristics of 3D point cloud data, we defined PA as the global proportion of correctly classified points across the entire point cloud. To account for potential volumetric imbalances among different anatomical structures, CPA was evaluated to reflect the prediction accuracy within each specific target class. Furthermore, spatial overlap was rigorously assessed using IoU—calculated as the ratio of the intersection to the union of the predicted and ground-truth points for a given class—with mIoU serving as the macro-averaged spatial congruence across all predefined ROI categories.

To estimate the statistical reliability of these evaluations, 95% confidence intervals (CIs) for all performance metrics were calculated using bootstrap resampling with 1,000 iterations, maintaining the original cohort class distributions. All statistical analyses and metric calculations were performed using Python (version 3.8) with the scikit-learn and SciPy libraries.

## Data availability

Due to patient privacy and institutional review board requirements, the raw three-dimensional imaging data and clinical records are not publicly available. All data requests supporting the results of this study should be directed to the corresponding author, J.L.Y..

## Acknowledgements

This work was supported by the National Key Research and Development Program of China (Research on prevention and treatment of common frequently-occurring diseases / 2023YFC2507702,2023YFC2507706)

## Author contributions

J.L.Y., L.A., and Y.Q.Z. conceived and supervised the project. J.F.Y., Z.H., T.Z., Z.Z., Z.F., X.X., Y.G., and Y.D. collected the original 3D imaging data and clinical records. J.F.Y. and Z.H. annotated the data. H.S., X.W., W.W., J.W., Y.Z., H.L., J.Z., and L.T. developed the algorithms and wrote the code. H.S. and X.W. performed the validation and statistical analysis. J.L.Y. and Y.Q.Z. provided clinical guidance and result calibration. L.W. and X.L. provided technical guidance. J.F.Y., H.S., Z.H., and X.W. wrote the manuscript. All authors discussed the results and commented on the manuscript.

## References

1. Zhang, Z. et al. Intelligent Scoliosis Screening and Diagnosis: A Survey.

2. Kim, H., Chang, B.-S. & Chang, S. Y. Current issues in the treatment of adolescent idiopathic scoliosis: a comprehensive narrative review. Asian Spine J. 18, 731–742 (2024).

3. Li, M., Nie, Q., Liu, J. & Jiang, Z. Prevalence of scoliosis in children and adolescents: a systematic review and meta-analysis. Front. Pediatr. 12, 1399049 (2024).

4. Kim, D. J. et al. The diagnostic accuracy of community spine radiology for adolescent idiopathic scoliosis brace candidates. Eur. Spine J. 33, 3776–3783 (2024).

5. Goldberg, C. J., Moore, D. P., Fogarty, E. E. & Dowling, F. E. Scoliosis: a review. Pediatr. Surg. Int. 24, 129–144 (2008).

6. Lai, K. K.-L. et al. Monitoring of Curve Progression in Patients with Adolescent Idiopathic Scoliosis Using 3-D Ultrasound. Ultrasound Med. Biol. 50, 384–393 (2024).

7. Chen, H. et al. Development of Automatic Assessment Framework for Spine Deformity Using Freehand 3-D Ultrasound Imaging System. IEEE Trans. Ultrason. Ferroelectr. Freq. Control 71, 408–422 (2024).

8. Zhu, X. et al. MGScoliosis: Multi-grained scoliosis detection with joint ordinal regression from natural image. Alex. Eng. J. 111, 329–340 (2025).

9. He, Z. et al. Conditional generative adversarial network-assisted system for radiation-free evaluation of scoliosis using a single smartphone photograph: a model development and validation study. eClinicalMedicine 75, 102779 (2024).

10. Ćuković, S. et al. Advancing 3D idiopathic scoliosis assessment through optical scans and web-based integration - Validation of the ScolioSIM system. Comput. Biol. Med. 196, 110735 (2025).

11. Mohamed, N. et al. Three-dimensional markerless surface topography approach with convolutional neural networks for adolescent idiopathic scoliosis screening. Sci. Rep. 15, 8728 (2025).

12. Zhou, Z. et al. Gait Patterns as Biomarkers: A Video-Based Approach for Classifying Scoliosis. in Medical Image Computing and Computer Assisted Intervention – MICCAI 2024 (eds Linguraru, M. G. et al.) vol. 15005 284–294 (Springer Nature Switzerland, Cham, 2024).

13. Dong, N., Zhang, X., Liu, X., Guo, W. & Wang, F. Key Points Positioning: A Two-Stage Algorithm For Single-view Point Cloud of Human Back Based on Point-wise Network. in Proceedings of the 2022 11th International Conference on Computing and Pattern Recognition 264–272 (ACM, Beijing China, 2022). doi:10.1145/3581807.3581846.

14. Rothstock, S., González-Ruiz, J. M., Weiss, H.-R., Turnbull, D. & Krueger, D. Severity and Cobb angle of scoliosis patients quantified with markerless trunk surface topography using k-NN search and multivariate regression analysis. Comput. Methods Biomech. Biomed. Eng. Imaging Vis. 11, 2433–2439 (2023).

15. Goldman, S. N. et al. Applications of artificial intelligence for adolescent idiopathic scoliosis: mapping the evidence. Spine Deform. 12, 1545–1570 (2024).

16. Amirian, S. et al. Explainable AI in Orthopedics: Challenges, Opportunities, and Prospects. Preprint at 10.48550/ARXIV.2308.04696 (2023).

17. Kadhim, M. et al. Current status of scoliosis school screening: targeted screening of underserved populations may be the solution. Public Health 178, 72–77 (2020).

18. Luan, F.-J., Wan, Y., Mak, K.-C., Ma, C.-J. & Wang, H.-Q. Cancer and mortality risks of patients with scoliosis from radiation exposure: a systematic review and meta-analysis. Eur. Spine J. 29, 3123–3134 (2020).

19. Lian, C. et al. Deep Multi-Scale Mesh Feature Learning for Automated Labeling of Raw Dental Surfaces From 3D Intraoral Scanners. IEEE Trans. Med. Imaging 39, 2440–2450 (2020).

20. Banerjee, A. et al. Point-Cloud Method for Automated 3D Coronary Tree Reconstruction From Multiple Non-Simultaneous Angiographic Projections. IEEE Trans. Med. Imaging 39, 1278–1290 (2020).

21. Hettikankanamage, N. et al. eXplainable Artificial Intelligence (XAI): A Systematic Review for Unveiling the Black Box Models and Their Relevance to Biomedical Imaging and Sensing. Sensors 25, 6649 (2025).

